# A Study of Patients undergoing Trigger Finger Release Surgery in a Primary Care setting – Organisational Capabilities, Outcomes and Benefits

**DOI:** 10.1101/2023.07.03.23292179

**Authors:** Fahad Rizvi, Chen Wei Rong Ryan, Kong Amos Ethan, Wong Chun Pui Joshua, Neal Khambhayata, Dhriti Arya, Tariq Kapasi, Philippe B. Wilson

## Abstract

**Background:** Trigger finger is a common hand condition in which a finger is unable to fully extend due to a thickening of the tendon and its sheath, causing the finger to lock in a bent position.

**Aim:** To assess the viability of carrying out Trigger Finger surgeries in NHS primary care in terms of clinician and patient acceptance, experience and outcomes, and operational requirements of this service for wider application.

**Design and Setting:** In this study, a total of 214 Trigger Finger Release Surgery procedures carried out between 22nd August 2019 and 25th October 2022 by a single hand surgeon in a single Primary Care surgery in Leicester, United Kingdom.

**Methods:** Data were analysed using data from SystemOne, which is a patient database linked with the National Health Service (NHS).

**Results and Conclusion:** Herein, we identify the opportunity to significantly reduce pressures on secondary care orthopaedic referrals as well as offer patients faster and effective surgical treatment within a primary care setting utilising far less NHS resources.

**How this fits in:** There are huge challenges in addressing long orthopaedic waiting lists in UK secondary care, therefore these can be alleviated by completing standard and non-complex cases by qualified surgeons in the primary care sector. We describe a clinic set-up including operational requirements and positive patient outcomes for Trigger Finger surgeries carried out within a primary care network in Leicester, UK.

## Introduction

Trigger finger is defined as tenosynovitis in the flexor sheaths of the fingers and thumb [1]. It can arise from repetitive gripping actions which can cause narrowing of the flexor pulley sheaths in the hands [2]. Progressively, the hypertrophy and inflammation of the retinacular sheath can restrict motion of the flexor tendons of the affected hand [3].

Trigger Finger can cause various impacts on quality of life [4], characterised by symptoms of the condition such as stiffness, locking and pain [5] which are frequently reported at the metacarpophalangeal joints [6].

Management can include conservative measures such as activity modification and analgesia for pain relief in the form of non-steroidal anti-inflammatory drugs (NSAIDs) [7]. However, steroid injections carried out under local anaesthetic are often recommended as the first-line treatment,[8] with referral for secondary care opinion if the patient expresses a preference for surgical release or if symptoms fail to resolve after steroid injections [9]. Indeed, Schubert and colleagues have described demographics of trigger finger occurrence and corticosteroid injection therapy as a retrospective review, identifying a significantly higher proportion of female cases (71%) versus 29% for males, with women reporting symptoms at an average age of 58 compared to 62 for men. Prevalence of diabetes in the cohort was 22% and no correlation was observed between trigger digit and dominance of hand, whilst the right index and thumbs were the most common;y-affected digits [10].

Surgery in the form of either an open or percutaneous release of the affected finger is the current recommended method to treat Trigger Finger [11]. This highly successful procedure is well recognised as the most effective treatment for Trigger Finger [12].

However, current waiting times for outpatient appointments in Leicestershire alone rest at 17 weeks for OPA and 22 weeks for treatment [13]. We describe a primary care service developed with an average patient waiting time of 4 weeks in comparison, as well as the operational and post-operative outcomes associated with the running of this service.

## Aims

This study aims to examine the clinical outcomes of Trigger Finger Release Surgeries conducted in a Primary Care setting in order to understand the impact of carrying out such procedures in the community. Furthermore, we detail the operational requirements and learnings from developing the service in order to provide other organisations with the opportunities to expand this offering into their own service distributions.

This study pays particular interest in establishing current waiting times from a patient’s first referral from their general practitioner (GP) to the first out-patient appointment (OPA) at the Trigger Finger Clinic, and the subsequent lead time from the first OPA at the Trigger Finger Clinic to the Trigger Finger Release Surgery itself.

The post-operative (post-op) outcomes of these procedures are also scrutinised by analysing a range of related parameters such as the percentage of post-operative complications, the nature of such complications (e.g.: pain, infection, swelling), and the follow-up actions required for each case.

## Methods

A total of 214 Trigger Finger Release Surgery procedures carried out between 22nd August 2019 and 25th October 2022 by a single hand surgeon in a single Primary Care surgery in Leicester, United Kingdom, were scrutinised using data from SystemOne, which is a patient database linked with the National Health Service (NHS). These constituted of procedures carried out on 107 Male and 107 Female patients. For each procedure, we recorded the hand operated on (left or right) and the finger of that hand that was operated on (thumb, index, middle, ring, little). The date of the patient’s first GP referral was identified, together with the date of the first OPA clinic with the hand surgeon. Prior to this, the patient has the opportunity to have a telephone discussion with their GP and the surgeon to discuss referral if they wish. Alternatively, the patient has the opportunity to be directly referred for their first OPA which thereby delivers the fast turnaround times for initial consultation and completion of the procedure. We also noted if there were any corticosteroid injections administered for each case, and if so, the date on which it was administered. Finally, the date of each Trigger Finger Release Surgery procedure was recorded.

Following this, we identified any post-op follow-ups on SystemOne. If no further data was identified after the date of the procedure, we assumed that there would be no patient or clinician initiated follow-up and the case was closed. However, if further data was found after the date of the procedure, then this would be marked as a follow up and checked for any post-op complication. We recorded the total number of post-op complications, and for each complication we noted the nature of the complication (*e*.*g*. pain, infection, swelling), the date that it was identified, and whether any follow-up action was taken (*e*.*g*. referral to Secondary Care, steroid injection, antibiotics, etc), and if so, the date of action taken.

## Results and Analysis

The average patient would spend 28 days waiting for an OPA clinic appointment with the hand surgeon (Table 1). This data is spread out with moderate consistency given the standard deviation of 27.8 days. It should be noted that 32 cases (15.0% of all cases) experienced a wait time of less than ten days from the GP referral to the 1st OPA, with two cases being seen on the same day by the hand surgeon as the GP referral itself. There were only six cases (less than 3% of all cases) which experienced a wait time of more than 100 days and this was attributed to patient choice of having the procedure at a later date due to their commitments. Essentially, this highlights the capacity for a quick referral from the GP to the 1st OPA with the hand surgeon, which could save precious time for patients.

**Table 1:** A summary of mean and median waiting times during the study.

|  |  |
| --- | --- |
| Mean time taken from GP referral to 1st OPA in days | 28.0 |
| Median time taken from GP referral to 1st OPA in days | 23.0 |
| Standard deviation (in days) of time taken from GP referral to 1st OPA | 27.8 |
| Mean time taken from 1st OPA to surgery in days | 51.3 |
| Median time taken from 1st OPA to surgery in days | 14.0 |
| Standard deviation (in days) of time taken from 1st OPA to surgery | 90.7 |

The data for the time taken from the 1st OPA clinic to the actual surgery itself was more dispersed, with a standard deviation of 90.7 days. Given the mean time from the 1st OPA to the surgery (51.3 days), the average person would likely wait less than two months prior to undergoing surgery after the 1st OPA clinic.

Given that standard practice guidance as described earlier indicates 2 injections prior to operation, patients had already experienced injections previously and indicated a preference for surgery. Therefore based on a combination of patient choice as well as clinical guidance, 39 cases had opted for an initial treatment of steroid injections at the 1st OPA before eventually opting for surgery. This could possibly have lengthened the time recorded from the 1st OPA to the surgery, thus affecting the obtained data. Furthermore, personal availability for the surgery is a contributing factor, as with the possibility that some patients could have preferred to see how symptoms developed prior to the surgery.

It should be highlighted that 71 cases (33.2% of all cases) experienced less than a ten day waiting time between the 1st OPA and the surgery itself, with 18 cases (8.41% of all cases) undergoing surgery on the same day as the 1st OPA itself. This was due to patients’ previous experience of trigger release, failed steroid treatment and opportunity to get the surgery on the same day, if not they are offered a trigger release procedure within 4 weeks. This suggests that there was a capacity for almost a third of all cases to be seen at a reasonably short notice, which could suggest the efficiency and time management advantages of conducting Trigger Finger Release Surgery in Primary Care.

Out of the total of 214 unique procedures carried out, there were 26 post-op complications that required follow-up action (Figure 1), which comprised 12.1% of the total number of procedures carried out. Of the total number of cases where post-op complications were identified, 13 cases required further follow-up action, while the remaining 13 did not have any identifiable further follow-up action, and were regarded as closed cases thereafter. 11 suspected post-op infections were identified, where signs and symptoms such as pain, swelling and the development of pus were reported. Out of all 214 cases performed, this equates to 5.14%. 12 cases were identified as depicting signs and symptoms suggestive of post-op mechanical symptoms (*e*.*g*. stiffness, locking). This equates to 5.61% of the total number of procedures performed, as shown in Figure 1.

**Figure 1:**
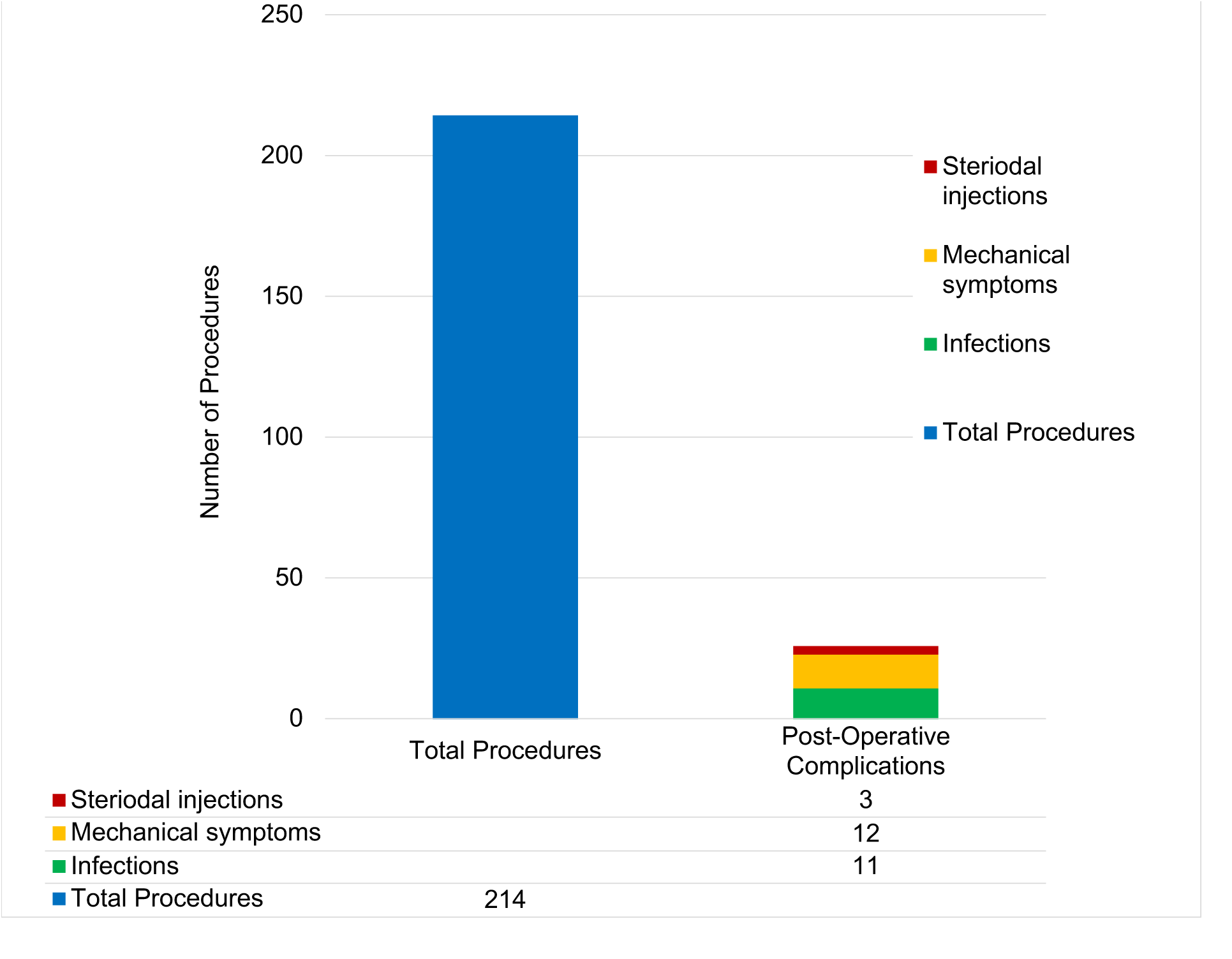
Classification of post-operative complications compared to total procedures carried out within the study. The label terms are refer to the following: Steroid injection for tenosynovitis following trigger release; Mechanical symptoms – stiffness, swelling and scar inflammation; Infection – superficial infection treated with normal dose(500mg qds for 7 days) flucloxacillin, or deep infection which required a surgical washout for tendon sheath infection. In this case series only 1 patient required surgical intervention.

Three cases required steroid injections after the procedure, which makes up 1.40% of all 214 cases. One case was identified as requiring further surgical intervention after the Trigger Finger Release Surgery. This represents 0.47% of all the 214 procedures performed.

This data suggests that there are reasonably low chances of post-op complications developing, with the vast majority of cases not requiring further post-op action. Given that complications for Trigger Finger Release Surgery can be as high as 31% [14], the significantly lower post-op complication rate of 12.1% in our study indicates a potential benefit of conducting Trigger Finger Release Surgery in Primary Care. Furthermore, a 5.7% risk of peripheral nerve damage with percutaneous release has been reported [14], hence many surgeons favouring open release as an alternative.

The number of operations carried on each hand (left or right) and the number of different fingers involved (*i*.*e*. thumb, index, middle, ring or little) are described in Table 2. 55% of procedures were carried out on the right hand, with the middle and ring fingers being the most likely fingers to be operated on. Conversely, the thumb was operated on in only 5% of cases.

**Table 2.** The number of operations carried out on the hand and the individual fingers.

|  | Left hand | Right hand |
| --- | --- | --- |
| Total on each hand | 96 | 118 |
| Thumb | 3 | 8 |
| Index | 11 | 9 |
| Middle | 39 | 45 |
| Ring | 36 | 44 |
| Little | 7 | 12 |

### Service Operationalisation and Delivery

#### Rationale

The service was designed to leverage the existing expertise in primary care and simultaneously aim to reduce waiting lists at the local acute trust. Indeed, a 2015 review by Morrissey and coworkers considered the potential as well as output impact of interdisciplinary triage and treat services at the primary care interface for musculoskeletal conditions [15]. Across the 23 studies identified, 72% of patients or above were able to be managed at an earlier stage through intermediate care pathways, leading directly to a 20-60% reduction in orthopaedic referral rates. Furthermore, there have been further references to such interfacial services for the reduction in orthopaedic outpatient waiting lists, specifically in consideration of the shifting policy landscape within the UK NHS [16]. Ferguson and Cooke (2011) assessed the NHS Bath and North East Somerset Orthopaedic Interface Service, where an intermediate service resulted in only 21% of patients being referred to secondary care, the remainder being successfully managed within primary care settings.

#### The setting and integration within the integrated care model

The referral areas was that of the Leicester City, Leicestershire and Rutland (LLR) CCG covering more than 1.2m patients. The caseload described within this study accounted for 50% of the cases, where the remaining 50% of referrals were undertaken by secondary care. All referrals were made by the Referral Service Scheme which is a standard NHS practise where this offering was integrated into the opportunity. Furthermore, patients were referred to our service as part of the standard clinical pathway including previous iterations of corticosteroid injections prior to surgical referral.

#### The approach

Our methodology is based on the concept of GP retraining where indeed a number of clinicians in primary care have sought additional training in specialty fields which they are able to leverage. In this instance, a GP with surgical specialism and qualifications collaborated with a local Consultant Hand Surgeon in the acute care trust in order to establish this service. The local GP surgical specialist and consultants working in the primary care setting were allocated 250 procedures per year in order to achieve the availability described.

Surgeries were carried out in existing, purpose-built minor surgery units in primary care, with more than or equal to 15 air changes per hour. The facility benefitted from a Service Level Agreement with the local Care Commissioning Group at the time. Operationally, the service was able to run at just above half the current NHS tariff for such procedures, as the economies of scale within the primary care units allowed for savings on porters, multiple theatre assistants, large administrative teams and estates/overhead costs. Indeed, the current NHS tariff to secondary care for such procedures is marginally over £1,000 per case, whereas the service described within this study was able to deliver such procedures at a 40% cost saving, thereby £600. In comparison, the team used to deliver this service consisted of: GP specialist or Consultant, theatre assistant (mid-band nurse), district nurse (for follow-ups) and an administrative colleague to arrange appointments and additional clerical tasks.

Patients were called at the surgery slot and in most cases would have been discharged from the surgery within 30 minutes of arrival, thereby avoiding unnecessary waits, overcrowding and patient anxiety. Follow up was carried out by the district nurse team for stitch removal and benfitted from open-access patient initiated follow-up appointments (PIFUs) for any concerns or queries relating to the service. This avoided patient anxiety concerning additional clinician contact and reduced likelihood of emergency department visits given the open-access nature of the PIFUs as an alternative.

#### Evaluation and post-operative care

Patients receive a hand evaluation questionnaire 6 weeks post-surgery in order to assess their recovery and capture any concerns, with the option of scheduling a follow-up appointment.

Upon patient initiated follow up, the surgeon would receive an automated email with any additional information such as wound photographs or patient notes and scheduled a slot to call the patient. This would be carried out as a same-day service either by audio or video call. The surgeon would discuss with the patient their preference and a face-to-face appointment set up if clinically required or requested by the patient.

The development of this service with demonstrable ease of access for patients, as well as the open and accessible follow-up opportunities in addition to the improved time and financial considerations indicate an overall benefit for implementation; indeed, of all the referrals triaged to primary care, locally 90% of trigger release interventions are carried out in primary care with 10% referred to the acute trust as the presentation was not commensurate to trigger finger, instead being a related condition such as Dupuytren’s Contracture, flexor or extensor injury, ganglions and arthritis.

## Discussion

### Summary

This study suggests potential benefits of conducting Trigger Finger Release Surgery in Primary Care with regards to waiting times and the quality of the procedures undertaken.

### Strengths and Limitations

Whilst this study was carried out as a pilot for a new service opportunity, the delivery and operationalisation of the service as well as patient outcomes suggest a strong opportunity for service extension to further primary care centres. Due to the nature of the service being based in a single primary care network, the number of participants being 214 may be considered relatively low, however this is pertinent when considered in the wider context of evaluating the delivery of this new service for potential extension to other community-based offerings.

### Comparison with Existing Literature

Such surgical interventions have been found to have success rates of over 90% in a recent study [12]. Moreover, over the 214 procedures undertaken, only 12.1% of these resulted in post-operative complications, significantly lower than the up to 31% notes previously [14]. Futhermore, of the 12.1% post-operative complications, only half of these required follow-up actions, arguably indicating that a total of 6% of all cases performed justified treatment. This suggests that risk factors and prevalence of such complications are not increased by undertaking prcoedures in a primary care setting, instead we see both an improvement in availability of such procedures to patients, with a marked improvement in post-operative complication rates. The ease of contacting the operating surgeon was readily available thus providing confidence to patients. The patients were provided phone numbers to contact without the need to return to their GPs or attending busy A&E departments in working hours. They had open access to follow ups and with a variety of same day consultation options such as phone, video call, e-mail or face to face as per the patients’ convenience and request.

### Implications for Research and Practice

A number of cases were identified and operated on the same day, with successful clinical outcomes such as low post-op complication rates. These are suggestive of the high efficiency and standards associated with this practice in Primary Care, which could contribute to overall patient satisfaction and their quality of life with early return to work. In the future, this practice has the potential to reduce Secondary Care pressures should more Trigger Finger Release Surgeries be managed in a Primary Care setting. Furthermore, procedures carried out in primary care, such as open release, can also allow for initial indentification of less common conditions such as traumatic or atraumatic ruptures of the flex digitorum profundus [17]. A 2019 study carried out at the University of Utah considered cost implications of surgical interventions in operating rooms *versus* those carried out in minor theatres or procedure clinics, providing a similar comparison between the secondary and primary care intervention environments in the UK. Principally, and considering the healthcare funding landscape differences in the US, the study identified a 221% cost increase from carrying out Trigger Finger Release Surgeries in operating theatres as opposed to procedural clinics [18]. The *British Society for the Surgery of the Hand* (BSSH) in their *Evidence for Surgical Treatment* brief [19] carried out a systematic review and meta-analysis of the following treatment options described in the literature: corticosteroid injections, open surgery and percutaneous release. Of the trials analysed, the 2001 study by Gilberts *et al*. followed percutaneous release and open surgery patients within 12 weeks of the procedure and did not identify any difference in recurrence rates which were low for both procedures [20]. Furthermore, a 2015 randomised study by Sato *et al*. compared open pulley release, percutaneous release and steroid injections in 150 digits over 6 month follow-ups in a population aged over 15 years. Whilst no complications were noted, equivalent outcomes in both percutaneous and open pulley release surgery were reported [21]. Sripheng and colleagues recently reported results of a 12-year retrospective observational study with a 2.39% recurrence rate over 841 fingers. More than 3 previous steroid injections and a history of manual labour were identified as independent predictors of recurrence [22].

Of the *Good Practice Points* identified in the BSSH report, recommendations to practitioners centred around referral to secondary care for surgeries. The study presented within the current manuscript suggests a real opportunity to relieve pressures on otheopaedic clinics in secondary care by offering such procedures in a primary care setting.

## Data Availability

All data produced in the present study are available upon reasonable request to the authors

## Ethical Approval

This work was undertaken as a service evaluation and therefore internal ethical approval was obtained with Reference: EB/14-40-2019.

## Acknowledgements

The authors thank the Partners at NHS Willows Health for supporting the activities described within this manuscript.

